# Epidemiological Assessment of Accidents and Functional Limitations among Nursing Home Residents in Shiraz, Iran (2024)

**DOI:** 10.64898/2026.01.30.26345186

**Authors:** Marzieh Sheibani, Hassan Rezaeipandari, Hossein Fallah

**Affiliations:** Elderly Health Research Center, Department of Aging and Health, School of Public Health, Shahid Sadoughi University of Medical Sciences, Yazd, Iran; Industrial Diseases Research Center, Department of Ergonomics, School of Public Health, Shahid Sadoughi University of Medical Sciences, Yazd, Iran

**Keywords:** Ergonomics, Accidental Falls, Nursing Homes, Aged, Accident Prevention

## Abstract

**Background:** With the rapid ageing of Iran’s population, accidents among institutionalised older adults represent a major public health concern. This study aimed to determine the prevalence, characteristics, and risk factors of accidents among elderly residents of nursing homes in Shiraz, Iran, during 2024, with particular emphasis on functional limitations.

**Methods:** A cross-sectional census-based study was conducted in all seven nursing homes in Shiraz, involving 550 residents aged ≥60 years. Data were collected through structured interviews, review of medical records, caregiver reports, and an Accident Form. Accidents occurring during the previous year were analyzed using descriptive statistics, and associations between accident occurrence and participant characteristics were examined using chi-square tests.

**Results:** Overall, 72.0% of residents experienced at least one accident during the study period. Slipping was the leading cause, and bathrooms and toilets were the most frequent locations. Contusion or bruising was the most common outcome. Mobility limitation was the only factor significantly associated with accident occurrence (p < 0.001), whereas age, gender, marital status, and educational level showed no significant associations.

**Conclusions:** Accidents were common among nursing home residents in Shiraz and were strongly associated with mobility limitation. These findings highlight the importance of addressing functional impairments alongside environmental hazards through targeted ergonomic modifications and mobility-support interventions.

## 1. Introduction

### 1.1 Background

Increased life expectancy and population aging represent one of the achievements of the 21st century, with population aging emerging as a phenomenon confronting certain human societies now or in the future [1]. It is projected that societal aging will persist over the next 50 years, accompanied by a rise in disease burden and associated costs [2, 3]. According to World Health Organization estimates, by 2050, Iran, as a developing country, will have a substantial elderly population [4]. Projections indicate that by 2050 (equivalent to 1430 in the Iranian calendar), Iran’s elderly population will reach approximately 19.92% of the total population [5].

### 1.2 Problem Statement

Given the economic, social, and chronic disease challenges faced by the elderly, along with the inability of most families to provide daily medical and nursing care, referrals to nursing homes have increased. Currently, more than 2 million people worldwide reside in nursing homes [6]. At present, approximately 230 elderly care centers operate in Iran; [7] thus, considering the unique circumstances of the elderly and their need for care and maintenance, the necessity for nursing homes is evident, and the quality of care provided in these facilities holds paramount importance [8].

Aging constitutes a stage of human life that naturally accompanies a decline in physical and mental abilities [9, 10] . As age advances and this period is reached, the functional abilities of the elderly diminish [11]. In this context, the unfavorable changes encountered by many elderly individuals pave the way for numerous problems [12]. Aging, through disruptions in motor function, is associated with reduced balance sensation and impaired performance of balance control systems—such as diminished strength, speed, sensory systems, coordination, and neural control—which in turn lead to decreased functional mobility, social and physiological issues, and ultimately an elevated risk of accidents [13]. With advancing age, sedentary behavior and motor poverty increase, thereby causing balance deficits and subsequent accidents among the elderly [14]. In this study, an accident refers to incidents occurring in the elderly’s living environment that result in identifiable injuries [8]. The five primary causes of accidents, which account for the highest incidence of home-based incidents among the elderly, include contact with hot liquids, contact with sharp objects, tripping, falls, and collisions with objects (impact) [13]. Based on conducted studies, the highest incidence of accidents and mishaps occurs among elderly individuals [15-17]. In a study in Mazandaran on elderly individuals admitted to pre-hospital emergency services, the highest accident rate (33.8%) occurred among the elderly, with priorities including traffic accidents (59.8%) and falls (32%) [18]. In the study by Khazaei et al., traffic accidents (32.74%), falls (21.49%), and impacts (20.49%) were reported as the most frequent accident occurrences among the elderly [19].

Approximately 40-80% of falls among elderly individuals living among the general community result in injuries. Falls in the elderly constitute the most common single factor limiting their activity and leading to immobility [20]. Scientific sources indicate that 27 to 65% of elderly individuals experience at least one fall [21]. This statistic, from research on community-dwelling elderly in the country, stands at 25%, while for elderly residing in national care centers, it is reported as 48.3% for women and 51.7% for men [22].

### 1.3 Study Rationale

According to reports from the United States Centers for Disease Control and Prevention, non-compliance with standards leads to accidents and injuries in elderly individuals. For instance, falls among elderly residents in nursing homes are twice as frequent as among those living in the community. The most common causes of falls in these facilities include wet sloped stairs, insufficient environmental lighting, inappropriate bed height, unsafe wheelchairs, and the absence of protective handrails near elderly beds [23]. In elderly living environments, particularly in Asian countries, no adaptations have been made to align physical and environmental conditions with their needs [24].

Attention to the elderly’s living environment represents a critical domain in their care. Given the diversity of environments in various elderly care centers, designs for elderly living in these facilities must incorporate ergonomic principles based on the functionality of components and objects present [25, 26]. Preventing accidents and mishaps in the places where the elderly reside can be achieved through environmental modifications. Such modifications enhance human-environment harmony and contribute to improved quality of life for the elderly [27, 28].

### 1.4 Objectives

Considering the evident and undeniable rapid growth of the elderly population in the country, and given that aging entails conditions including disabilities, neglecting them could lead to severe negative consequences. In this regard, the objective of this research is to determine the prevalence of accidents among elderly residents in 24-hour care centers in Shiraz.

## 2. Methods

### 2.1 Study Design

This study employed a cross-sectional descriptive-analytical design conducted in 2024 to examine the epidemiology of accidents among elderly residents of nursing homes in Shiraz, Iran. The research was field-based and quantitative, relying on structured data collection from institutionalised older adults rather than laboratory experimentation.

### 2.2 Participants

The target population comprised all older adults aged 60 years and above residing in the seven public nursing homes supervised by the Welfare Organization of Fars Province, Shiraz, Iran. A complete census approach was adopted, eliminating sampling bias and yielding a final sample of 550 residents. Inclusion criteria were permanent residency in the facility, age ≥60 years, and provision of informed consent (by the resident or legal guardian when cognitive impairment precluded self-consent). Residents who declined participation or were hospitalised throughout the data-collection period were excluded.

### 2.3 Data Collection

Data were collected using two standardised instruments. First, a researcher-designed demographic and clinical questionnaire recorded age, sex, marital status, education, duration of residency, chronic diseases, medication use, and functional limitations. Second, accident events during 2024 were documented via a structured incident registration form that captured cause, location, timing, associated activity, and clinical outcome. All tools had undergone content validity assessment by a panel of gerontology and ergonomic experts. Data collection occurred between July and August 2024 through face-to-face interviews with residents, review of official nursing-home incident logs, consultation with caregivers. Each accident was cross-verified from at least two sources to minimise recall bias. Participant recruitment and data collection were conducted between 1 July 2024 and 31 August 2024.

### 2.4 Data Analysis

Data were entered into SPSS version 27. Descriptive statistics (frequencies, percentages, means ± standard deviations) and inferential tests (χ^2^ and Fisher’s exact test) were performed with statistical significance set at p < 0.05.

### 2.5 Ethical Considerations

Ethical approval for this study was obtained from the Research Ethics Committee of Shahid Sadoughi University of Medical Sciences, Yazd, Iran (Approval ID: IR.SSU.SPH.REC.1403.079). Written informed consent was obtained from all participants. In cases where cognitive impairment precluded the provision of informed consent, consent was obtained from the participants’ legal guardians. All data were collected anonymously and used solely for research purposes.

## 3. Results

### 3.1 Participant Characteristics

A total of 550 elderly residents from seven public nursing homes in Shiraz participated in this census-based study. During the year 2024, 396 residents (72.0%) experienced at least one accident, confirming a high incidence of accidental injury in institutional settings. The majority of incidents were non-fatal, with contusion being the predominant outcome. Mobility limitation emerged as the only participant characteristic significantly associated with accident occurrence.

The descriptive statistics of the characteristics of the older adults under study are presented in Table 1. Mobility limitation was the most prevalent functional restriction (58.0%), followed by hearing impairment (24.2%) (Table 2).

**Table 1.**
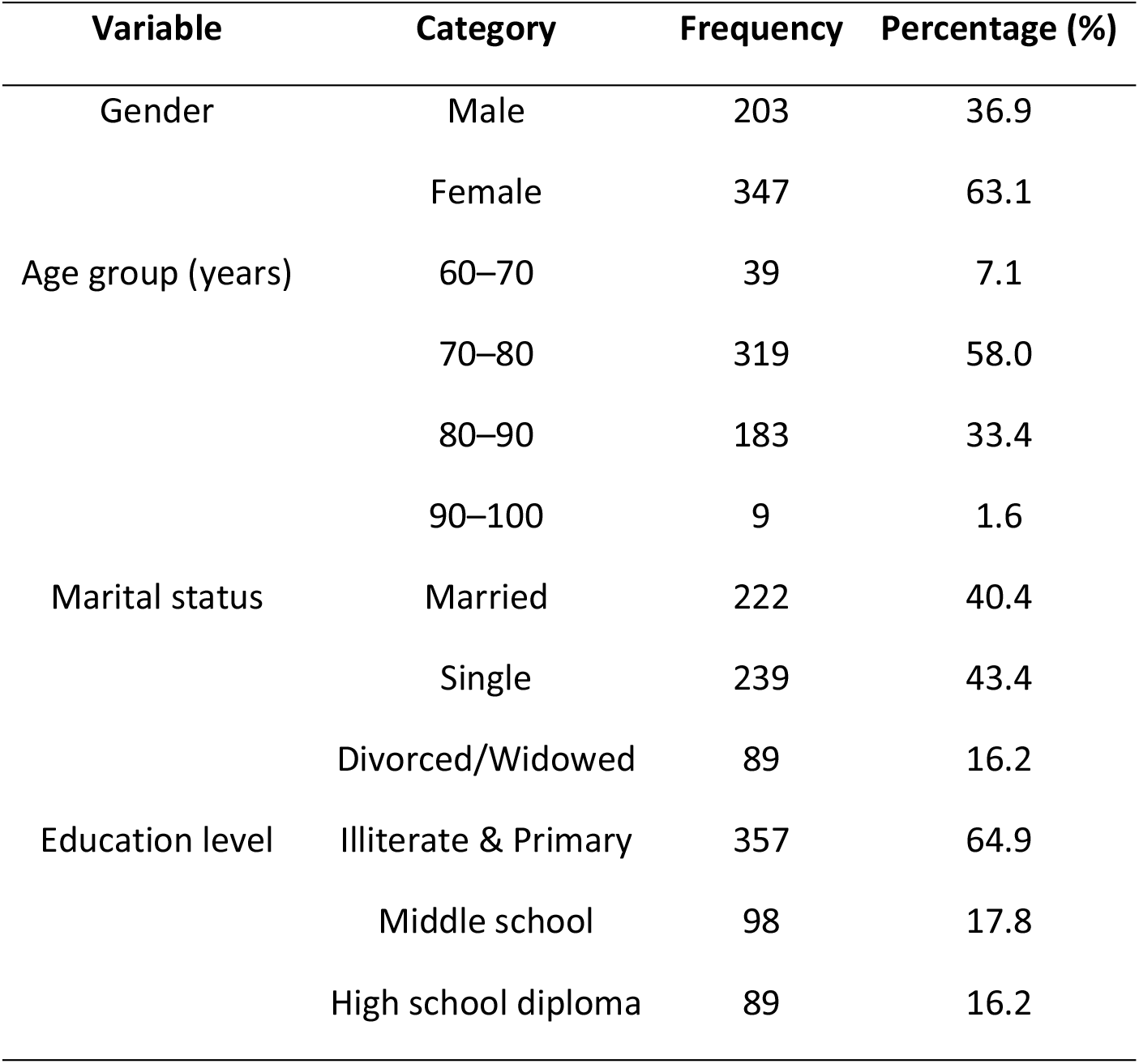
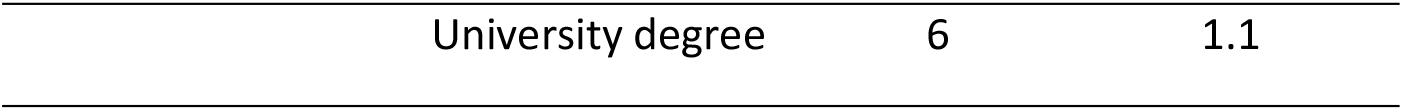
Descriptive statistics of qualitative demographic variables of the participants.

**Table 2.** Frequency distribution of limitations among the elderly.

| Variable | Response | Frequency | Percentage (%) |
| --- | --- | --- | --- |
| Mobility limitation | Yes | 319 | 58.0 |
|  | No | 231 | 42.0 |
| Hearing limitation | Yes | 133 | 24.2 |
|  | No | 417 | 75.8 |
| Visual limitation | Yes | 77 | 14.0 |
|  | No | 473 | 86.0 |
| Memory limitation | Yes | 112 | 20.4 |
|  | No | 438 | 79.6 |

### 3.2 Epidemiology of Accidents

No statistically significant associations were observed between accident occurrence and demographic variables, including age group, gender, marital status, educational level, or duration of residency (all χ^2^ p-values > 0.05; Table 3).

**Table 3.**
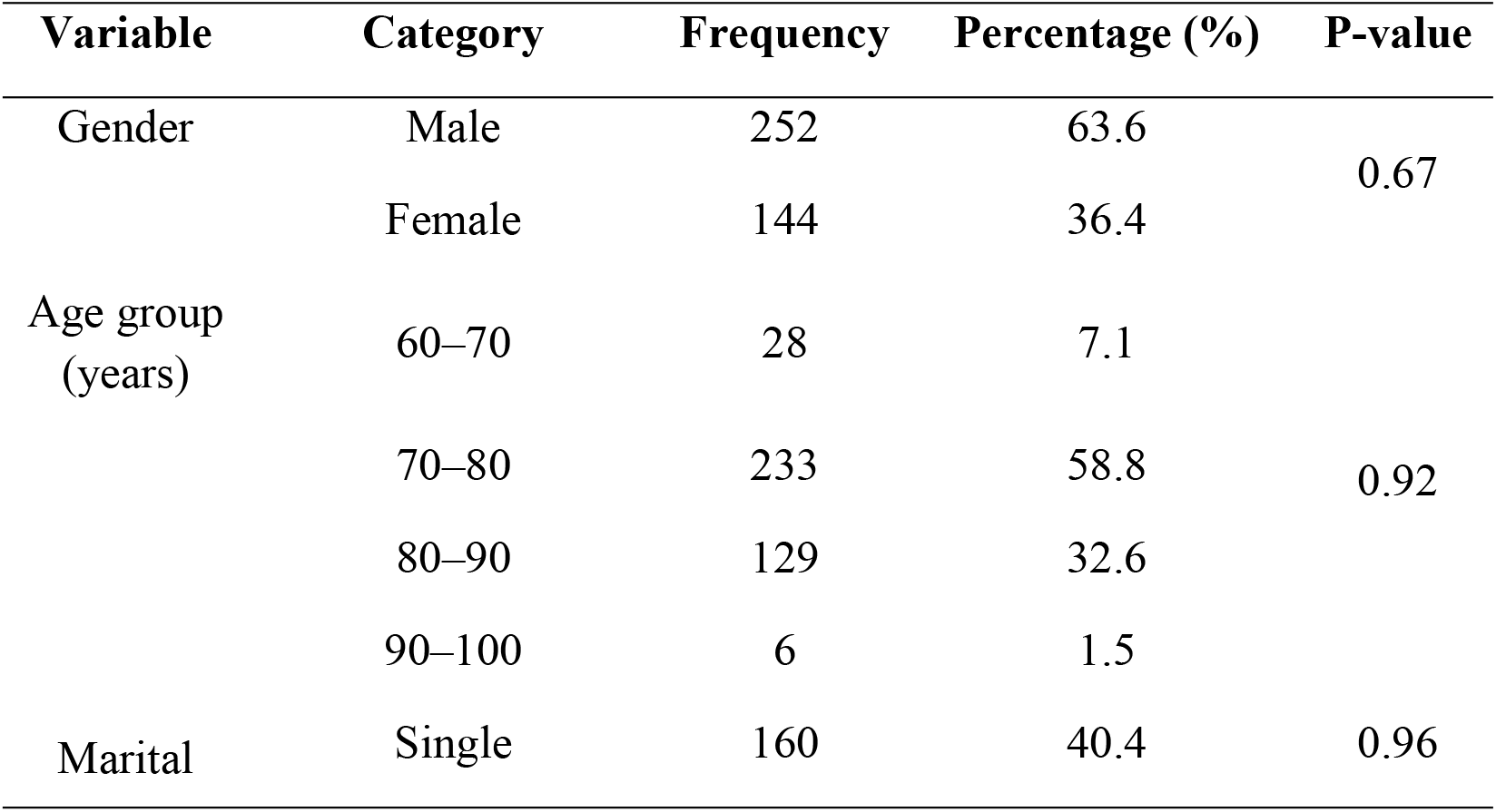
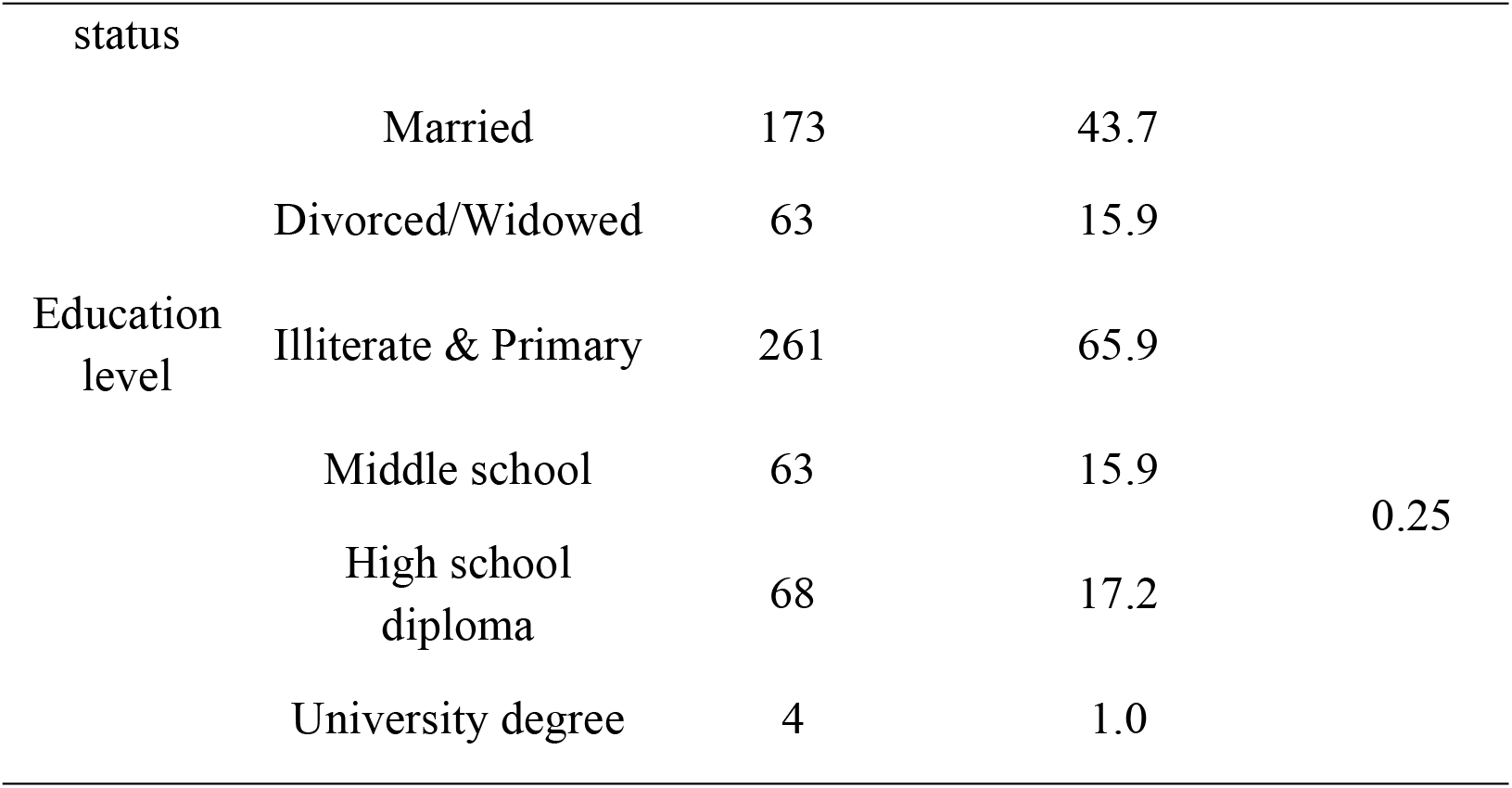
Frequency distribution of Accidents occurrence according to demographic variables.

Slipping was identified as the predominant cause of accidents, followed by collisions with objects and tripping. Incidents occurred most frequently in bathroom and toilet areas, with bedrooms and corridors ranking next. Toileting-related activities constituted the most common circumstance at the time of accident, whereas walking and transfers from bed or chair were less frequent. Contusion or bruising was the most prevalent injury outcome, while fractures and other injuries occurred less often (Table 4).

**Table 4.**
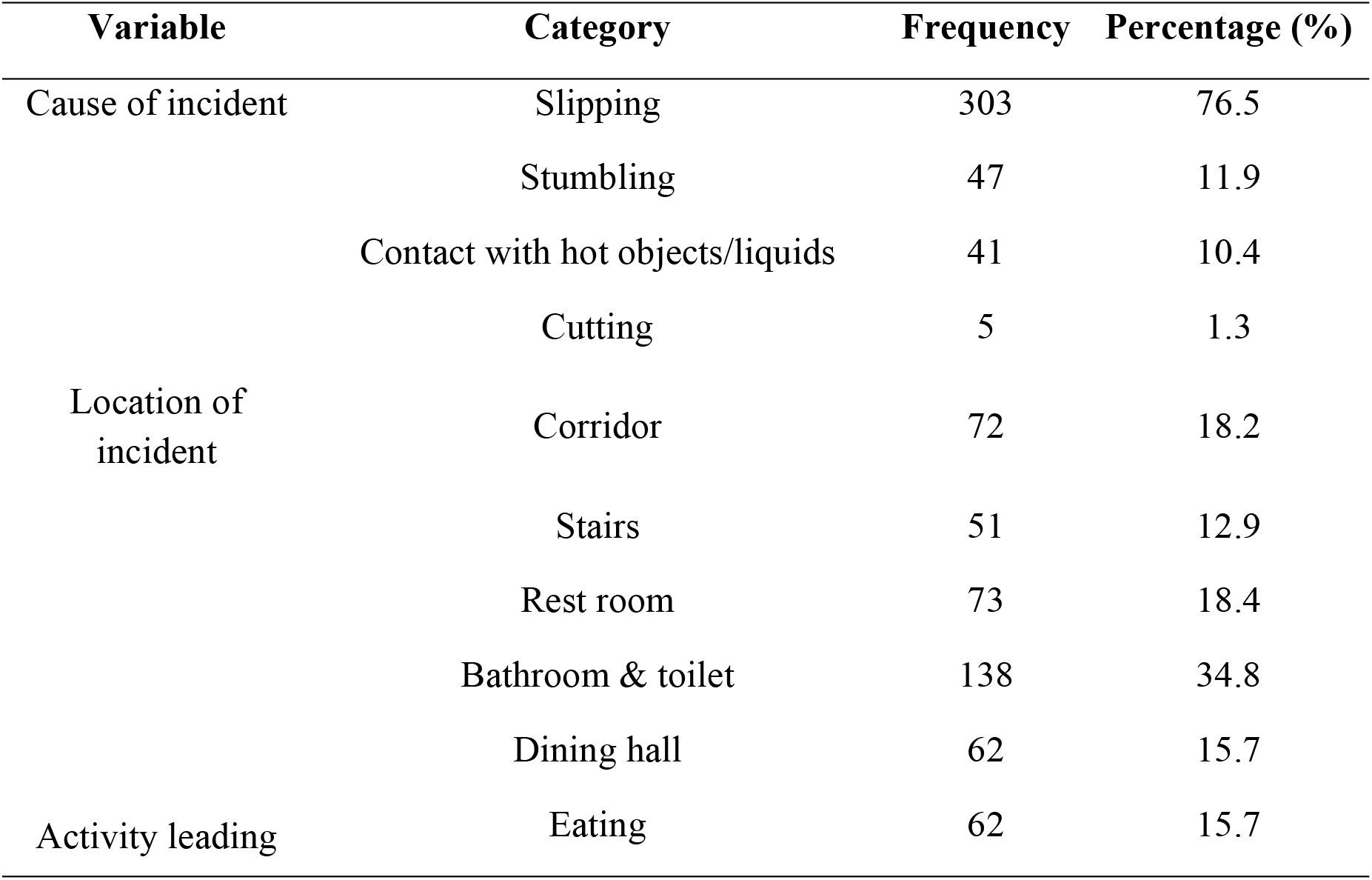
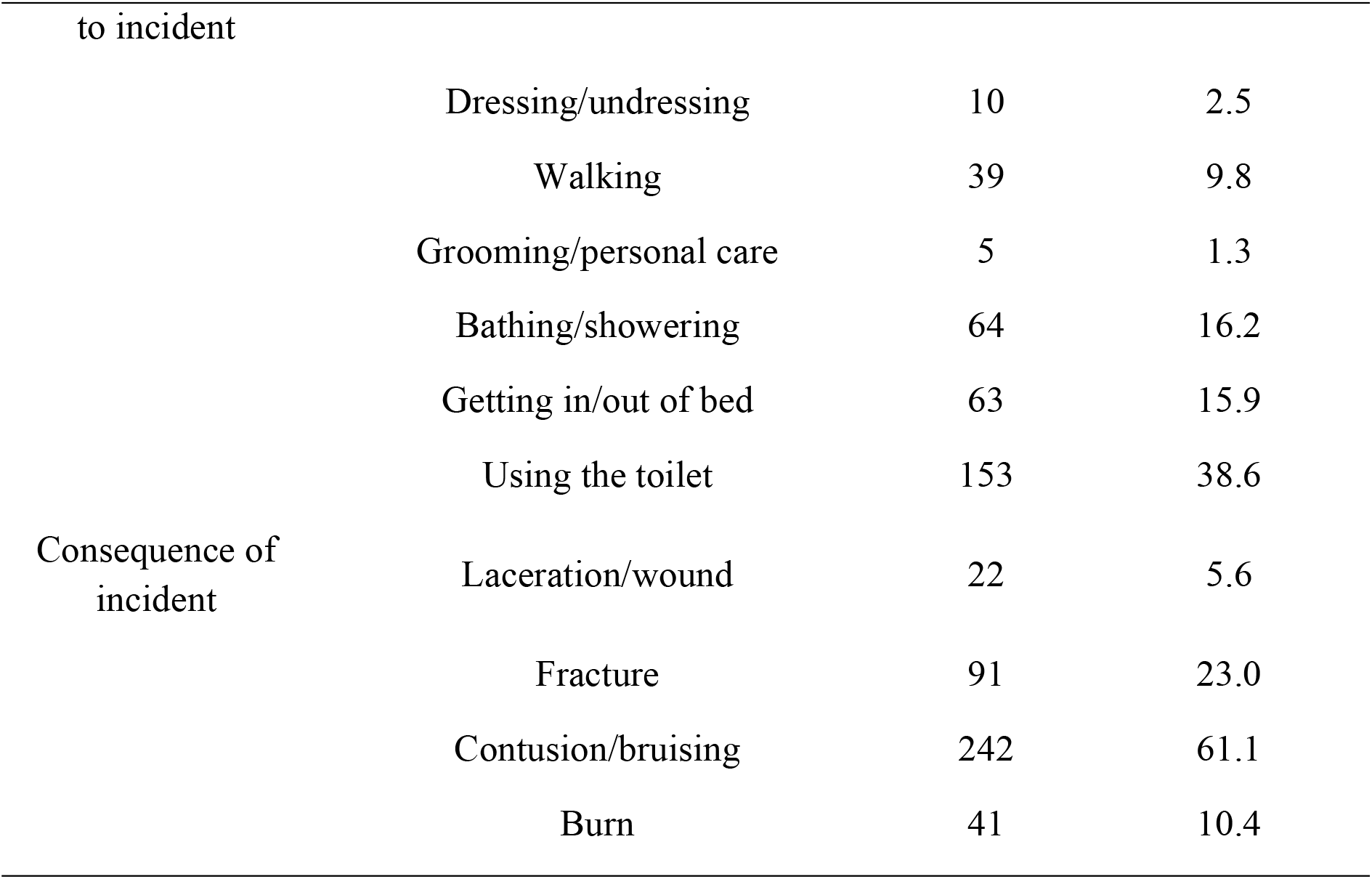
Frequency distribution of cause, location, activity, and consequence of incidents.

### 3.3 Association Between Limitations and Accident Occurrence

Mobility limitation demonstrated a strong and statistically significant association with accident occurrence (χ^2^ = 12.47, p < 0.001). Among residents with mobility limitation, 82.4% experienced at least one accident compared to 59.2% of those without limitation (Table 5). No other functional limitations (visual, hearing, or cognitive) reached statistical significance in relation to accident history. These findings indicate that impaired mobility was the primary intrinsic factor contributing to elevated accident risk in this institutionalised population.

**Table 5.** Association between elderly limitations and history of Accident.

| Variable | Response | Frequency | Percentage (%) | P-value |
| --- | --- | --- | --- | --- |
| Mobility limitation | Yes | 283 | 71.5 | <0.001 |
|  | No | 113 | 28.5 |  |
| Hearing limitation | Yes | 101 | 25.5 | 0.24 |
|  | No | 295 | 74.5 |  |
| Visual limitation | Yes | 55 | 13.9 | 0.90 |
|  | No | 341 | 86.1 |  |
| Memory impairment | Yes | 74 | 18.7 | 0.11 |
|  | No | 322 | 81.3 |  |

## 4. Discussion

### 4.1 Interpretation in Context of Existing Literature

The present study revealed a one-year accident prevalence of 72% among institutionalised older adults in Shiraz. The particularly strong association between mobility limitation and accident occurrence (p < 0.001) indicates that reduced lower-limb strength, gait instability, and dependence on assistive devices dramatically increase vulnerability when residents encounter common environmental hazards such as wet floors or absence of grab bars [29].

This interaction explains why simple activities like toileting became the most frequent triggers, transforming routine daily tasks into high-risk events and highlighting the urgent need for environmental modifications that compensate for age-related functional decline [30].

Compared with previous Iranian research, the 72% prevalence reported here substantially exceeds rates observed among community-dwelling elderly and aligns more closely with institutional studies elsewhere that document annual fall rates of 50–70% [31, 32].

The predominance of bathroom-related incidents mirrors international evidence identifying this location as the highest-risk zone in nursing homes due to combined slippery surfaces and limited supportive fixtures [30, 33].

However, unlike several community-based studies that identified female gender or advanced age as significant predictors, no demographic variables reached statistical significance in the present institutional sample, suggesting that extreme frailty and environmental exposure may override traditional risk profiles in nursing-home settings [34].

These results extend earlier Iranian work limited to single facilities or structural standards by providing the first census-based epidemiological profile for an entire metropolitan area [31].

### 4.2 Limitations of the Study

There are methodological limitations that require consideration. The cross-sectional design did not allow for the determination of the order of events or causality between the factors of limited mobility and accidents, and having to depend on the retrospective reporting of incidents during the year introduced memory bias, especially for the residents with cognitive impairments. The research was conducted only in public nursing homes in Shiraz, so its findings cannot be applied to private establishments or other areas with different standards of infrastructure. Furthermore, detailed information about the socioeconomic status and complete medication lists were not gathered, hence, control of possible confounding factors such as polypharmacy was not possible.

### 4.3 Practical Implications

It is suggested that future long-term cohort studies be carried out that involve objective evaluations of gait and balance, the monitoring of incidents in real-time, and multicenter sampling throughout Iran in order to verify causal pathways, as well as to assess the efficacy of environmental interventions like non-slip flooring and strategically placed grab bars with respect to the target groups [32, 35, 36].

## 5. Conclusion

### 5.1 Summary of Major Outcomes

Researchers conducted a census-based study involving 550 elderly residents from all public nursing homes in Shiraz, Iran, and found that 72% of the elderly people had at least one accident during the year 2024, with slipping being the leading cause and the bathroom/toilet area the most dangerous. Mobility limitation was the only factor significantly associated with accident occurrence. This finding suggests that accident risk increases when functional impairments interact with environmental hazards. The results address the objectives of the study and provide a detailed epidemiological profile of accidents in public nursing homes in Shiraz, Iran.

### 5.2 Implications for Policy and Practice

The implications of the findings outlined in this report are significant for immediate integration into geriatric health care systems and safety in facilities for older persons within Iran and other middle-income countries. The study suggests that cost-effective and evidence-based changes to the physical environment of care settings for older adults\textendash such as adding slip-resistant floors, installing grab bars in proper locations, improving the quality of lighting, and ensuring accessible toilets\textendash are key interventions which would lower the risk of injury and enhance the ability of residents with limited mobility to independently live; This should therefore lead to the creation of National Policy for Ergonomics in Nursing Facilities throughout Iran and other middle-income countries [37-39].

### 5.3 Recommendations for Future Research

Future research should evaluate the effectiveness of such modifications through controlled intervention trials and extend longitudinal monitoring to establish causal pathways and long-term outcomes. Expanding multicentre studies to include private facilities and rural regions will further enhance generalisability and inform nationwide prevention strategies.

## Declarations

### Data Availability Statement

The data underlying the results presented in this study contain sensitive information about elderly nursing home residents, including details on functional limitations, accident history, and demographic characteristics. Due to ethical restrictions and to protect participant privacy, as approved by the Research Ethics Committee of Shahid Sadoughi University of Medical Sciences, Yazd (code: IR.SSU.SPH.REC.1403.079), the full dataset cannot be made publicly available. De-identified data are available from the corresponding author upon reasonable request and subject to approval by the ethics committee.

## Acknowledgements

The authors thank the management and staff of the public nursing homes in Shiraz and the participating residents and caregivers. AI-assisted tools were used for language editing only.

## Authors’ Contributions

Conceptualization: Marzieh Sheibani, Hassan Rezaeipandari, Hossein Fallah.

Data curation: Marzieh Sheibani.

Formal analysis: Marzieh Sheibani, Hossein Fallah.

Investigation: Marzieh Sheibani.

Methodology: Marzieh Sheibani, Hassan Rezaeipandari, Hossein Fallah.

Supervision: Hassan Rezaeipandari, Hossein Fallah.

Validation: Marzieh Sheibani, Hossein Fallah.

Writing – original draft: Marzieh Sheibani.

Writing – review & editing: Marzieh Sheibani, Hassan Rezaeipandari, Hossein Fallah.

All authors have read and agreed to the published version of the manuscript.

